# HIV co-infection increases the risk of post-tuberculosis mortality among persons who initiated treatment for drug-resistant tuberculosis

**DOI:** 10.1101/2023.05.19.23290190

**Authors:** Argita D. Salindri, Maia Kipiani, Nino Lomtadze, Nestani Tukvadze, Zaza Avaliani, Henry M. Blumberg, Katherine E. Masyn, Richard B. Rothenberg, Russell R. Kempker, Matthew J. Magee

## Abstract

Little is known regarding the relationship between common comorbidities in persons with tuberculosis (TB) (including human immunodeficiency virus [HIV], diabetes, and hepatitis C virus [HCV]) with post-TB mortality. We conducted a retrospective cohort study among persons who initiated treatment for rifampicin-resistant and multi/extensively drug-resistant (RR and M/XDR) TB reported to the country of Georgia’s TB surveillance during 2009-2017. Exposures included HIV serologic status, diabetes, and HCV status. Our outcome was all-cause post-TB mortality determined by cross-validating vital status with Georgia’s death registry through November 2019. We estimated adjusted hazard rate ratios (aHR) and 95% confidence intervals (CI) of post-TB mortality among participants with and without comorbidities using cause-specific hazard regressions. Among 1032 eligible participants, 34 (3.3%) died during treatment and 87 (8.7%) died post-TB treatment. Among those who died post-TB treatment, the median time to death was 21 months (interquartile range 7–39) post-TB treatment. After adjusting for confounders, the hazard rates of post-TB mortality were higher among participants with HIV co-infection (aHR=3.74, 95%CI 1.77–7.91) compared to those without HIV co-infection. In our cohort, post-TB mortality occurred most commonly in the first three years post-TB treatment. Linkage to care for common TB comorbidities post-treatment may reduce post-TB mortality rates.

## INTRODUCTION

Tuberculosis (TB) remains a leading cause of infectious disease death with an estimated mortality rate of nearly 1.6 million persons worldwide annually.^1^ The END TB strategy aims to reduce TB mortality from 2015 to 2035 by 95%.^2^ Importantly, the reported number of TB deaths is likely underestimated because TB-attributable mortality after treatment is not typically measured or reported.^3^ More importantly, WHO reported that TB related mortality consistently increased for the first time after more than a decade since 2020, likely due to the COVID-19 pandemic.^1^ To meet the END TB goals, it is essential to better understand TB-related mortality including those that occur after the completion of TB treatment.

Emerging epidemiologic evidence suggests that individuals formerly treated for TB disease may have higher mortality rates compared to the general population.^3,4^ Despite microbial cure and favorable clinical outcomes, individuals formerly treated for TB disease may have residual sequelae including pulmonary impairment or increased risk of non-communicable metabolic diseases.^5-8^ However, to date, little is known regarding the relationship between pre-existing comorbidities (i.e., comorbidities diagnosed prior to or at the beginning of TB treatment) and post-TB mortality. To substantially reduce TB-related mortality, it is necessary to identify host risk factors, including pre-existing comorbidities, that are associated with post-TB mortality.

Previous studies have identified several demographic and behavioral characteristics associated with increased risks of post TB-mortality.^4,9-13^ For example, highly drug-resistant forms of TB such as multidrug-resistant TB (MDR TB) are associated with higher rates of mortality post-TB treatment.^9,14,15^ However, the effect of common pre-existing comorbidities such as human immunodeficiency virus (HIV) co-infection, hyperglycemia (i.e., elevated blood glucose with or without diabetes), diabetes, or hepatitis C virus (HCV) co-infection remains unclear. Understanding the extent to which these pre-existing comorbidities impact post-TB mortality rates will help identify prevention strategies to reduce post-TB mortality rates.

Given existing knowledge gaps, we aimed to a) estimate the rate of all-cause mortality post-TB treatment and b) determine the association between pre-existing comorbidities and post-TB treatment mortality rates using data of persons who initiated treatment with second line TB drugs (SLDs) from the country of Georgia.

## RESULTS

### Study Population and Characteristics

During the study period, there were 5,385 RR and M/XDR TB treatment episodes recorded in the national TB surveillance system, 31.5% (1,697/5,385) of which were classified as newly diagnosed RR and M/XDR TB (i.e., no prior history of TB treatment) (Figure 1). Of these, 1,416 (83.4%) unique case met study eligibility criteria and were cross-referenced with the country of Georgia’s vital registry. We excluded 384 individuals without unique national identifier, leaving 1,032 individuals (72.9%) included in our analyses (Figure 1). Individuals excluded due to missing vital status were similar to individuals included in the analyses in regard to age, gender, prevalence of MDR or pre-XDR TB, cavitary disease, hyperglycemia, and hepatitis C and HIV co-infections. Among individuals included, the majority were male (73%), and median age was 35 years (interquartile range [IQR] 26 – 49) (Table 1). The prevalence of MDR TB was 44.7% (461/1,032) and pre-XDR TB was 29.1% (300/1,032). HIV co-infection was reported among 3.8% (39/1,032, 95% confidence interval [CI] 2.7 – 5.1) and the prevalence of any hyperglycemia was 22.7% (234/1,032, 95%CI 20.2 – 25.3), 115 (49.1% [115/234], 95%CI 42.8 – 55.5) of which were determined to have diabetes. HCV co-infection was reported among 14.7% (153/1,032, 95%CI 12.8 – 17.1) of study participants.

**Figure 1.**
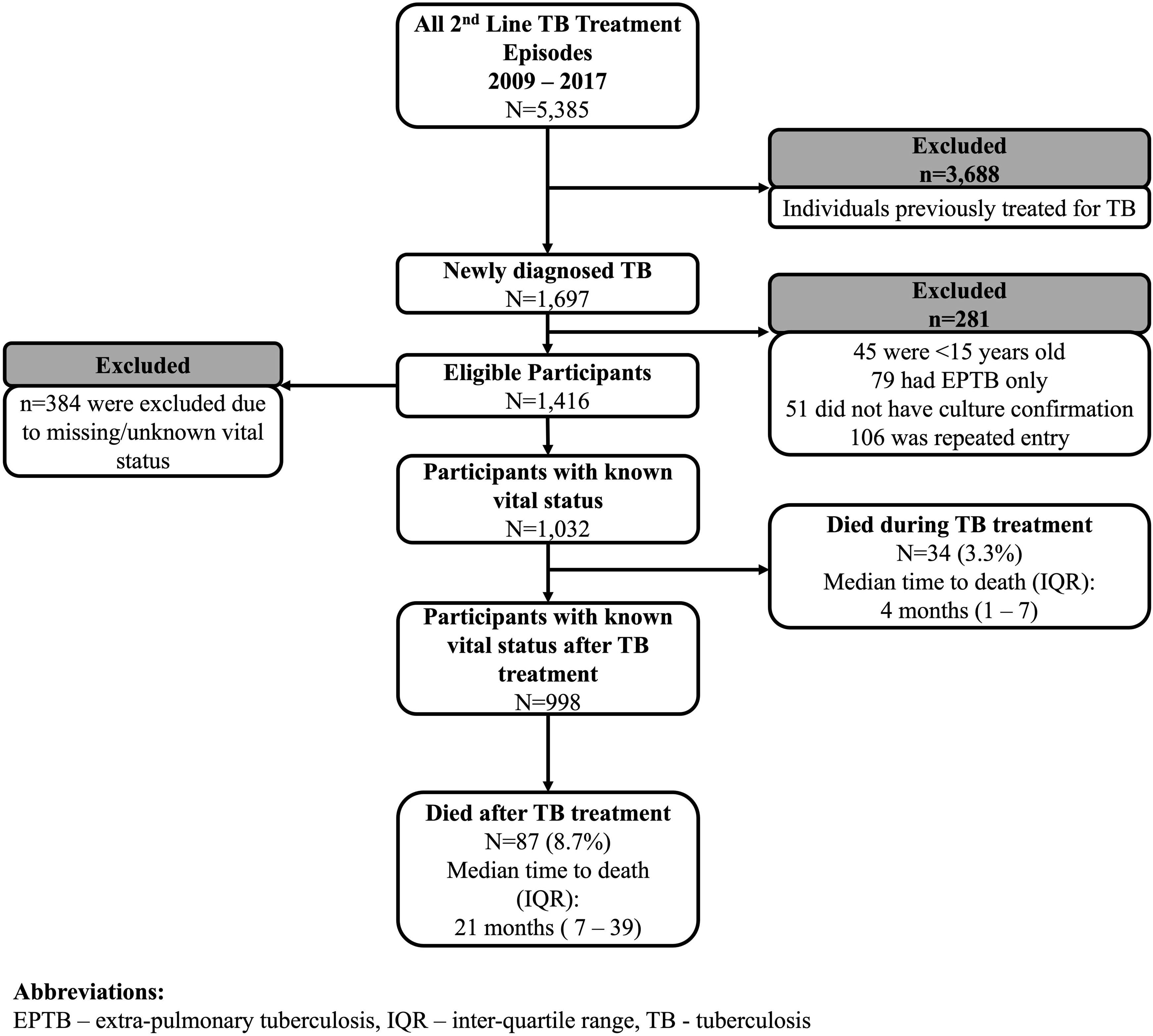
Participants selection according to inclusion criteria. This flowchart depicts the participants’ selection according to inclusion criteria (i.e., non-pediatric TB individuals with no prior history of TB treatment). Individuals were excluded if they had no pulmonary involvement, culture confirmation at the beginning of TB treatment, or unknown vital status during and after TB treatment.

**Table 1.**
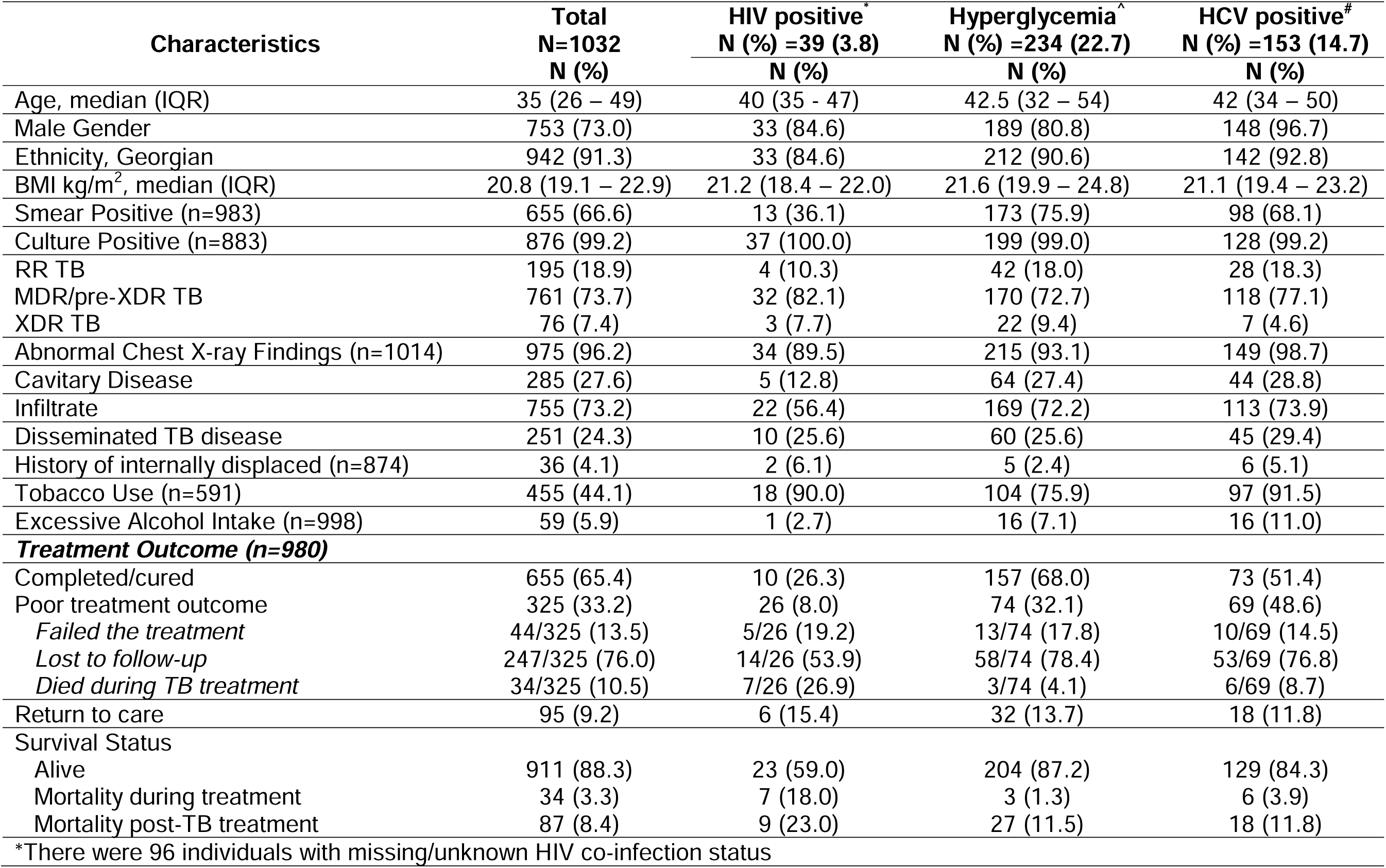
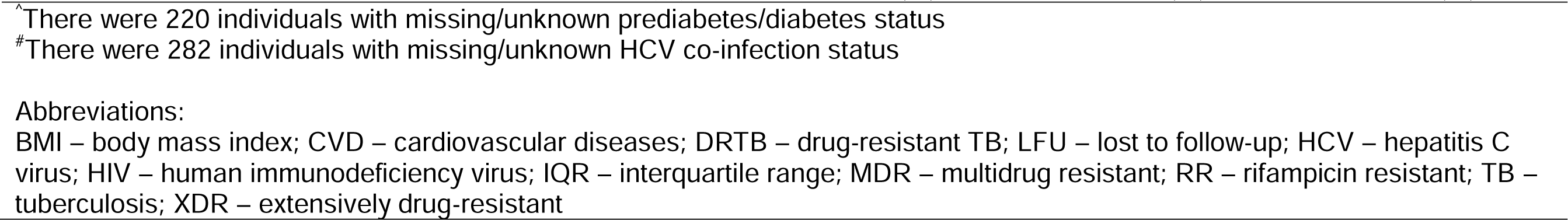
Demographic and clinical characteristics of persons who initiated treatment for RR and M/XDR TB according to pre-existing comorbidity status in Georgia, 2009 – 2017 (N=1,032)

Among 1,032 individuals included in the final analyses, 34 (3.3%) died during TB treatment during 1502 PY follow-up (age-adjusted mortality rate during TB treatment=22.6 per 1,000 PY, 95%CI 17.9 – 27.4) with a median time to death of 4 months (IQR 1 – 7) after TB treatment initiation. There were 87 (8.7%) post-TB deaths among 998 individuals alive at the end of their TB treatment during 4,857 PY of follow up (age-adjusted post-TB mortality rate=17.9 per 1,000 PY, 95%CI 11.9 – 23.9) (Table 2). Among those who died during TB treatment, the median age at time of death was 48 years (IQR 34 – 52). Nearly half (38/87, 43.7% 95%CI 33.6 – 54.2) of post-TB deaths were observed during the first year after TB treatment end, and the majority (67/87, 77.0%) occurred within the first three years with a median time to death of 21 months (IQR 7 – 39) after TB treatment was completed and/or stopped. Among those who died after-TB treatment, the median age at time of death was 50 years (IQR 39 – 63).

**Table 2.**
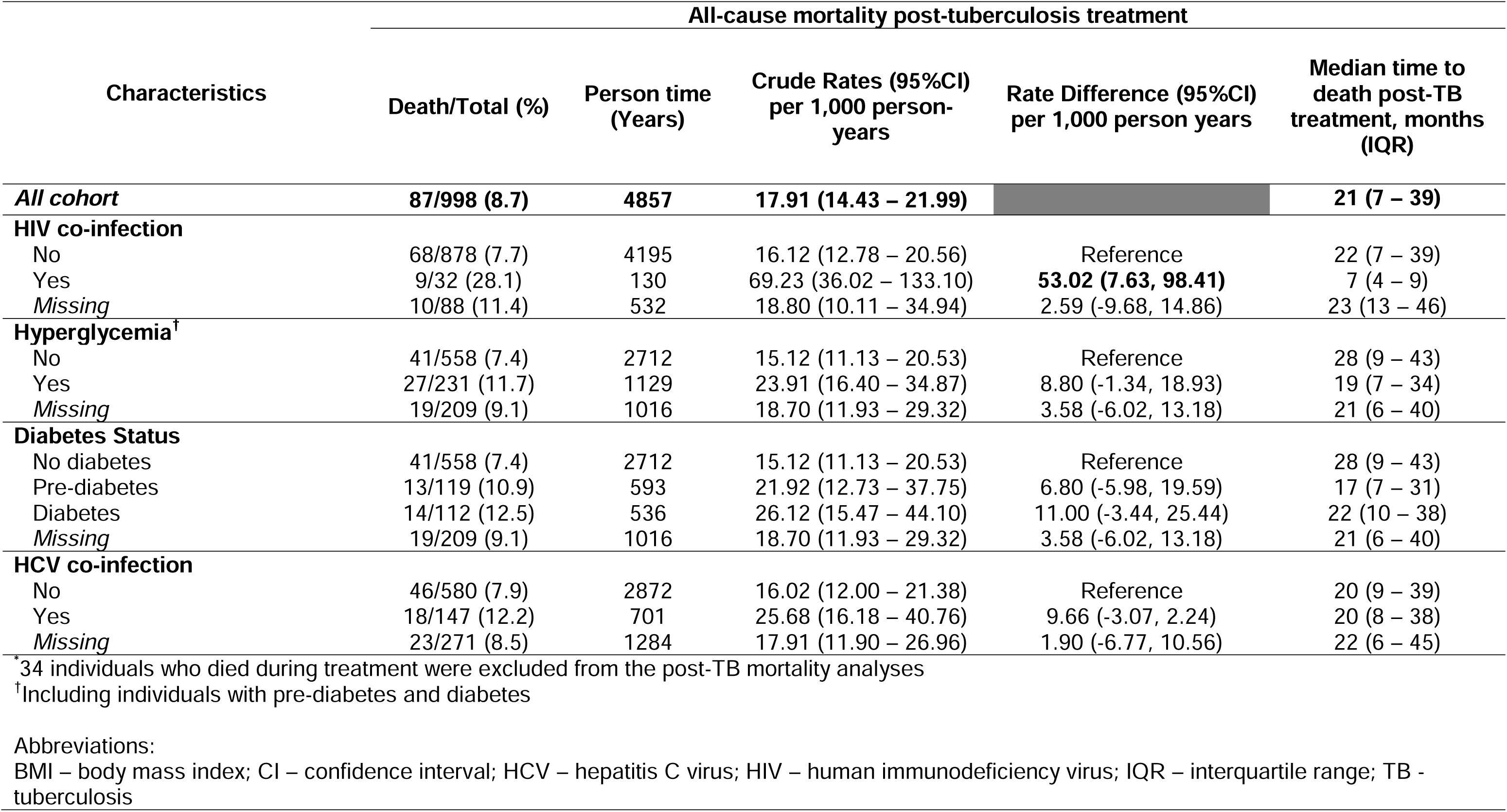
All-cause post-tuberculosis mortality rates among persons who initiated treatment for RR and M/XDR TB with and without pre-existing comorbidities in Georgia, 2009 – 2017 (N=998)*

### Pre-Existing Comorbidities and All-Cause Mortality Post-Tuberculosis Treatment

Among 32 persons living with HIV co-infection, there were 9 deaths post-TB treatment during 130 person-years (post-TB mortality rate 69.23, 95%CI 36.02 – 133.10). The median age at time of post-TB death among those with HIV co-infection was 41 years (IQR 39 – 46). The post-TB mortality rates among those with pre-diabetes was 21.92 (95%CI 12.73 – 37.75) and 26.12 (95%CI 15.47 – 44.10) among those with diabetes. The median age at time of post-TB death among those with pre-diabetes was 48 years (IQR 35 – 59) and 63 years (IQR 49 – 68) among those with diabetes. Among 147 individuals with HCV co-infection, there were 18 deaths post-TB treatment during 701 person-years (post-TB mortality rate 25.68, 95%CI 16.18 – 40.76). The median age at time of post-TB death among those with HCV co-infection was 57 years (IQR 39 – 62). Among 641 individuals without HIV, hyperglycemia, or HCV co-infection (i.e., no primary comorbidity), there were 41 deaths post-TB treatment during 3128 person-years (post-TB mortality rate 13.11, 95%CI 9.65 – 17.80) with the median age at time of post-TB death of 49 years (IQR 38 – 59).

Persons living with HIV co-infection had significantly higher hazard rate of post-TB mortality (adjusted hazard rate ratio [aHR]=3.74, 95%CI 1.77 – 7.91) compared to those without HIV co-infection after adjusting for potential confounders (Table 3). In the same adjusted model, the hazard rate of post-TB mortality among TB survivors with prediabetes was 1.48 (95%CI 0.78 – 2.78) and 1.06 (95%CI 0.53 – 2.12) among those with diabetes compared to those with euglycemic. The hazard rate of all-cause post-TB mortality among TB survivors with HCV co-infection was 1.21 (95%CI 0.68 – 2.59) times the hazard rate among those without HCV co-infection. The adjusted hazard rate of all-cause post-TB mortality among TB survivors with any primary comorbidity (i.e., HIV, prediabetes/diabetes, and HCV) was significantly higher (aHR=2.03 (95%CI 1.33 – 3.10) compared to those without. Our estimates did not change substantially in models with different covariates specification (Supplementary Table S1). We also provided post-TB mortality rates (Supplementary Table S2) and estimated hazard rate ratios (Supplementary Table S3) of post-TB mortality for other key risk factors (included other comorbidities, smoking status, and baseline BMI).

**Table 3.**
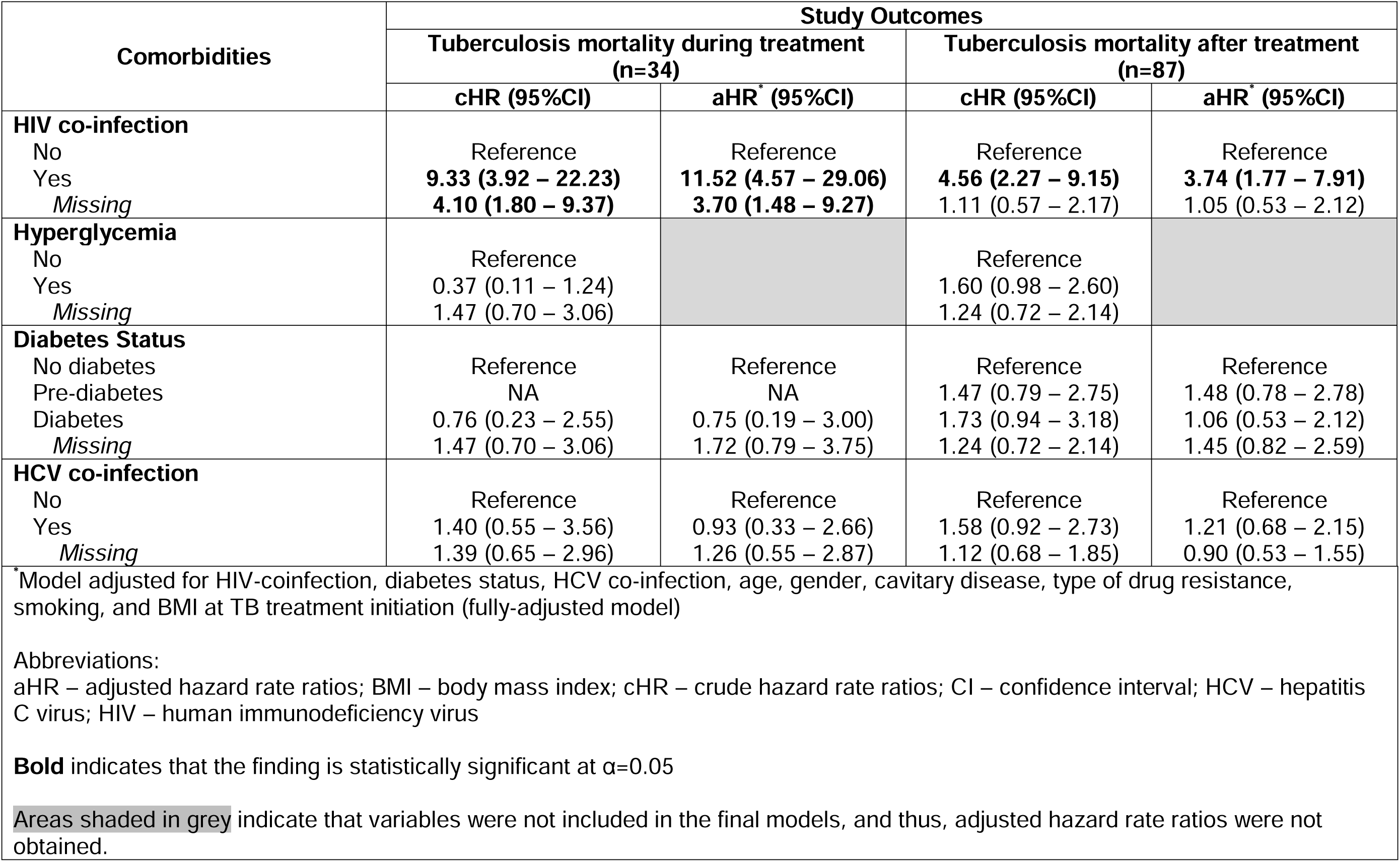
Crude and adjusted hazard rate ratios of mortality during and after tuberculosis treatment among persons who initiated treatment for RR and M/XDR TB with and without pre-existing comorbidities in Georgia, 2009 – 2017 (N=1,032)

### Results from Subgroup, Interaction, and Sensitivity Analyses

Among individuals with known smoking status (n=591), we found that the multiplicative effects of HIV co-infection and smoking, hyperglycemia and smoking, or HCV and smoking on mortality post-TB treatment were non-significant (statistical interaction p>0.05) (Table 4). We found that post-TB mortality rates were highest at 6-month post TB treatment among those with HIV co-infection, pre-diabetes/diabetes or HCV (Figure 2). Among those with HIV co-infection, pre-diabetes/diabetes, or HCV co-infection, the median time from the end of TB treatment to post-TB deaths were lowest among those aged 16 – 40 years (median=13 month, IQR 5 – 22) and ≥65 years (median=14 month, IQR 14 – 25) compared to those among age 41 – 65 years (median=27 month, IQR 6 – 39). Among those with known final TB treatment end (n=980), the hazard rate ratios of mortality post-TB treatment were similar among persons living with HIV or pre-diabetes/diabetes regardless of their final TB treatment outcome (Supplementary Table S4). However, those with HCV co-infection who were successfully treated for TB were more likely to die post-TB treatment compared to those with HCV co-infection but had poor final TB treatment outcomes.

**Figure 2.**
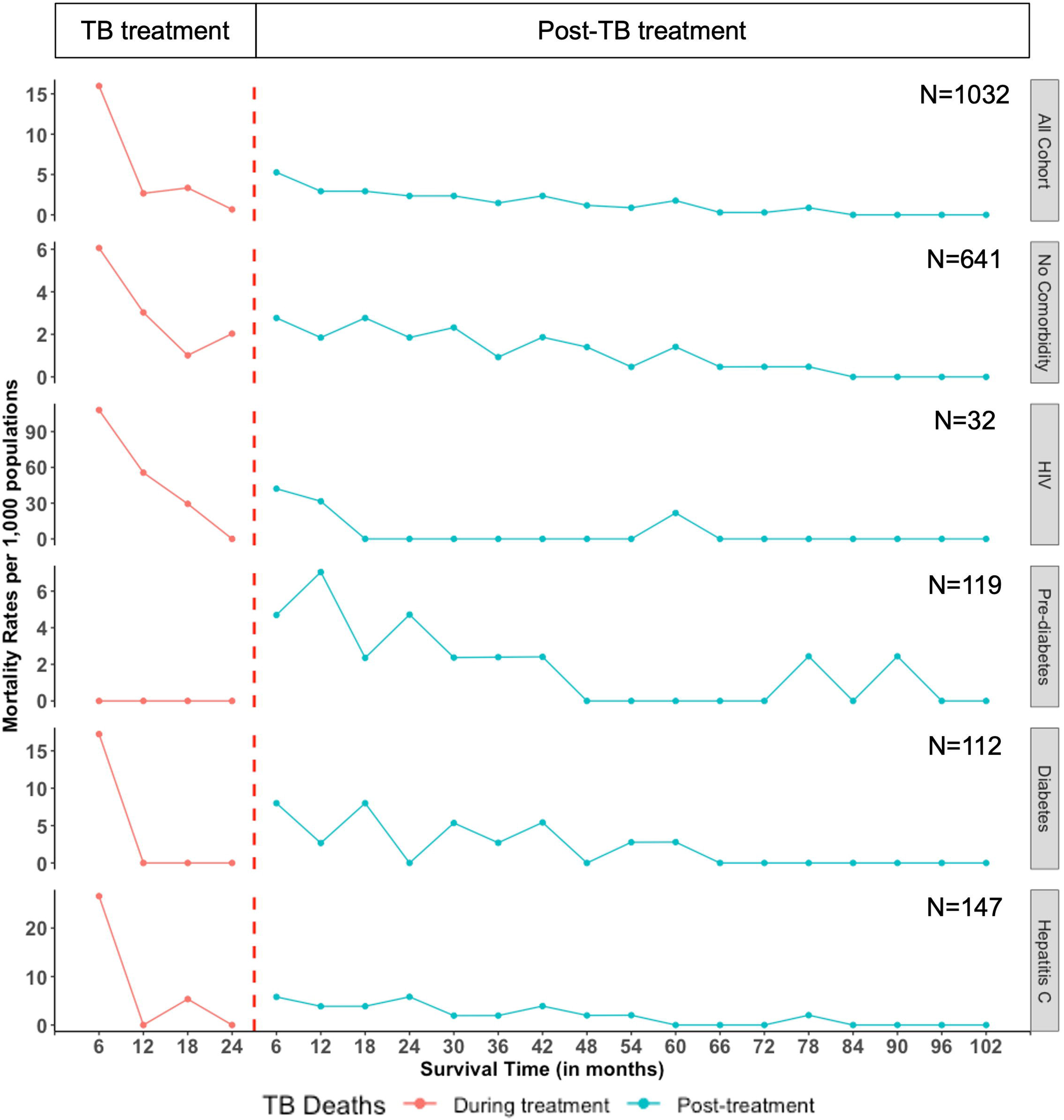
Trend of mortality rates reported during and after TB treatment stratified by comorbidity status among persons who initiated treatment for RR or M/XDR TB in the country of Georgia from 2009 – 2017. Mortality rates during and post-TB treatment were estimated by 6-month period by building independent life tables. Mortality rates during and post-TB treatment were expressed per 1,000 populations. Mortality rates were then plotted continuously with a single interruption point to mark the end of TB treatment (red dashed line). The beginning of survival time of ‘TB treatment phase’ (T_0-treatment_) marks the TB treatment initiation date. Thus, the point at T_6_ in the ‘TB treatment’ phase indicates the mortality rate at 6-month after TB treatment initiation. The beginning of survival time of ‘Post-TB treatment’ phase (T_0-post-TB treatment_) marks the end of TB treatment (i.e., could be treatment completion or lost to follow-up dates). Thus, the point at T_6_ in the ‘Post-TB treatment’ phase indicates the mortality rate at 6-month after TB treatment ended.

**Table 4.**
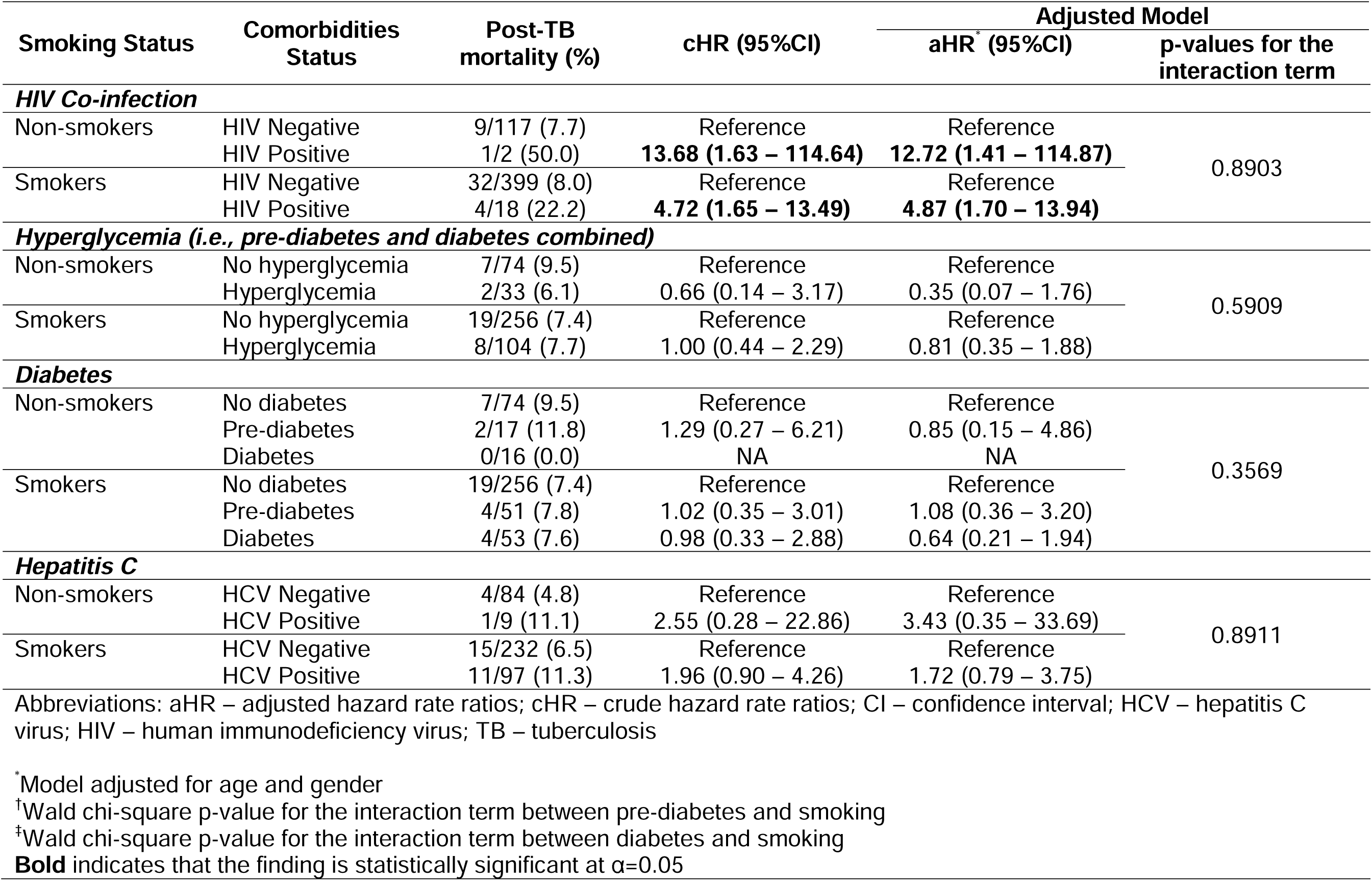
Subgroup analyses to assess statistical interaction between pre-existing comorbidities and smoking on mortality post-tuberculosis among persons who initiated treatment for RR and M/XDR TB in Georgia, 2009 – 2017 (N=591)

In the log-binomial models to estimate the cumulative risk of post-TB mortality, the cumulative risk of post-TB mortality among those living with HIV co-infection was 3.63 (95%CI 1.99 – 6.61) times the risk of those without HIV co-infection. The cumulative risk of post-TB mortality, although non-significant, was modestly higher among those with hyperglycemia (cRR 1.59, 95%CI 1.00 – 2.52) and HCV co-infection (cRR 1.53, 95%CI 0.92 – 2.58) (Supplementary Table S5).

The median of bias-adjusted RRs after accounting for any hyperglycemia misclassification was 1.84 (2.5^th^ – 97.5^th^ percentile 1.10 – 2.92) in the non-differential model vs. 1.73 (2.5^th^ – 97.5^th^ percentile 1.21 – 2.37) in the differential models. This indicates that misclassification of any hyperglycemia in our cohort may result in bias towards the null. The E-values of ≥6.72, ≥2.56, and ≥2.45 could explain away the observed association for HIV co-infection, hyperglycemia, HCV, and post-TB mortality risk.

## METHODS

### Study Design and Setting

We conducted a retrospective cohort study among adults with (>15 years old) newly diagnosed and laboratory-confirmed pulmonary TB. Persons initiated treatment with SLDs (including rifampicin-resistant and multidrug/extensively drug-resistant TB [RR and M/XDR TB]) and reported to the Georgian National Center for Tuberculosis and Lung Disease (NCTLD) surveillance system from January 2009 – December 2017 were eligible. Persons with drug-susceptible TB, persons with drug-resistant TB where strains were susceptible to rifampicin, and those with a history of prior TB treatment were excluded from analyses. We limited our analyses to those with newly diagnosed TB disease as Georgia’s online TB surveillance developed overtime and collection of prior TB information may not be uniform throughout our study period. More importantly, we believe that the underlying TB susceptibility profile of individuals with newly diagnosed TB may be different than those with a history of TB disease, thus, affecting the counterfactual conditions. Briefly, per Georgian National TB guidelines, standard of care performed at TB treatment initiation for those treated with SLDs includes complete blood count, chest X-ray, sputum smear and culture, drug susceptibility testing (DST), fasting plasma glucose (FPG), and rapid test of HIV infection. Additional tests to screen for other chronic comorbidities like viral hepatitis (hepatitis C and B) infections were done since 2008.

### Definitions and Study Measures

The primary exposures of this study were comorbidities diagnosed before or at TB treatment initiation, including a) HIV co-infection, b) any hyperglycemia, and c) HCV co-infection. HIV and HCV virus (HCV) co-infections were determined by antibody test performed at the beginning of TB treatment and recorded in the surveillance system. Any hyperglycemia was determined by either a self-reported previous diagnosis of type-2 diabetes (T2DM), or by FPG. Individuals with FPG ≥5.6 mmol/L or a self-reported prior T2DM diagnosis were categorized as “any hyperglycemia.” Individuals with FPG level <5.6mmol/L and no history of diabetes diagnosis were classified as “non-hyperglycemic.” We then separately defined diabetes status using three different levels: 1) no diabetes (FPG < 5.6 mmol/L with no previous diagnosis of T2DM), 2) pre-diabetes (FPG 5.6 – 6.9 mmol/L with no previous diagnosis of T2DM), and 3) diabetes (FPG ≥7.0 mmol/L or previous diagnosis of T2DM). “Any primary comorbidity” was defined if individuals had a record of either HIV co-infection, any hyperglycemia, or HCV co-infection. Other comorbidities recorded in the surveillance system (including self-reported cardiovascular disease, kidney disease, ulcer, pancreatitis, chronic obstructive pulmonary disease, silicosis, and sarcoidosis) were considered in the analyses by summing the number of comorbidities and classified as “none”, “1 – 2”, and “≥3”.

Our primary study outcome, all-cause mortality post-TB treatment, was determined by cross-referencing individuals’ national unique identifier with mortality status recorded in the death registry managed by the National Statistics Office of Georgia. Drug resistance was determined according to phenotypic DST results and were classified as rifampicin-resistant (RR), multidrug (MDR), pre-extensively (pre-XDR), and extensively drug-resistant (XDR) TB according to WHO guidelines.^16^ Other covariates included in our study (e.g., clinical TB treatment outcomes, self-reported smoking status, anthropometric measures) were defined according to surveillance records.

### Statistical Analyses

Chi-square/Fisher’s exact and Wilcoxon rank-sum tests were performed to assess associations between individuals’ comorbidities status and post-TB mortality (p-values were not shown). Poisson regression was used to estimate rates of all-cause mortality post-TB treatment (per 1,000 person-years [PY]) and rate differences (using identity link). We also built life tables to independently estimate the mortality rates per 6-month follow-up period during and after TB treatment. These mortality rates were then plotted with *ggplot2* package ^17^ in R Statistical Software (v4.2.3; R Core Team 2023) ^18^ to observe mortality trend during and after TB treatment. Proportional hazard models with competing risks were used to estimate the hazard rate ratios (HR) of all-cause mortality during and post-TB treatment comparing individuals with and without pre-existing comorbidities.^19^ Survival time was calculated from TB treatment initiation to a) time of death as recorded in the online TB surveillance system (for those who died during TB treatment), b) date of death recorded in the national death registry (for those who died after TB treatment), or c) the end of follow-up time for those who did not have the study events (i.e., alive by November 13, 2019, when mortality queries were cross-referenced with the vital registry’s office). In the cause-specific models estimating the HR of mortality post-TB treatment, individuals were censored if they a) died during TB treatment (i.e., competing risk) or b) had no death indication on the date mortality status was verified. Proportional hazard assumptions were assessed using 1) Kolmogorov-type supremum and 2) Schoenfeld’s residuals tests.^20^ Covariates included in the final multivariable models were selected considering established risk factors identified in previous studies, observed bivariate associations, and directed acyclic graphs.^21^

Stratified analyses were performed to determine whether the effect of pre-existing comorbidities on all-cause mortality post-TB treatment varied among those who had favorable (n=655) vs. poor TB treatment outcomes (i.e., including those who refused to continue treatment or were lost to follow-up) (n=325). Additionally, we performed a subgroup analysis to assess the interactions between pre-existing comorbidities and smoking status to determine if the association between pre-existing comorbidities and all-cause mortality post-TB treatment varied by smoking status. We assessed statistical interaction by including the cross-product terms (e.g., diabetes X smoking, HIV X smoking) in multivariable models. Statistical analyses were performed using SAS version 9.4 (Cary, North Carolina).

### Sensitivity Analyses

Sensitivity analyses were performed to quantify systematic errors (i.e., bias) due to a) distribution specification in the regression analyses, b) misclassification of any hyperglycemia status and c) confounding effects from unmeasured confounders. To assess distribution specification, log-binomial models were used to estimate cumulative risks of post-TB mortality comparing patients with comorbidities to those without comorbidities. Bias due to misclassification of hyperglycemia was of concern as we did not have a way to determine whether patients followed doctors’ instruction to fast at least 8 hours before test. To quantify bias due to hyperglycemia misclassification, we performed probabilistic bias analyses using beta distribution. To quantify systematic errors due to unmeasured/unknown confounders, we calculated E-values. E-value is an estimate of the minimum strength of association between unmeasured/unknown confounders with study exposures as well as study outcome to explain away the observed association between pre-existing comorbidities and all-cause mortality post-TB treatment.^22^

### Ethics Declaration and Approval

This study was submitted to, reviewed, and approved by the Institutional Review Boards (IRBs) at Georgia State University, Emory University, Atlanta, USA and the ethics committee at National Center for Tuberculosis and Lung Diseases, Tbilisi, Georgia (FWA00020831). Waiver of informed consent was obtained as data collected for this study were collected for TB surveillance purposes.

## DISCUSSION

In our large cohort of persons who initiated treatment with SLDs (n=1,032 with a median of follow-up time of 5 years), nearly 1 among 11 died post-TB treatment. We reported a higher rate of all-cause mortality rate in our cohort of persons treated for RR and M/XDR when compared to mortality rates of Georgia’s general population in 2019 (17.3 vs. 12.5/1,000 person-years).^23^ This increased rate of post-TB mortality is consistent with previously published studies in different settings.^3,4,10,14,24-26^ Furthermore, we found that post-TB mortality occurred mostly in the first three years after TB treatment ended. Additionally, we observed a significantly increased post-TB mortality risk among persons living with HIV co-infection compared to those without HIV co-infection. Understanding the effect of pre-existing comorbidities (especially HIV co-infection) on post-TB mortality could help determine whether additional care post-TB treatment completion is needed to reduce premature deaths associated with TB and comorbidities.

In our cohort, we reported nearly 4-fold increased hazard rates of all-cause mortality post-TB treatment among persons with RR and M/XDR and HIV co-infection compared to those without. However, this finding needs to be interpreted with caution as the number of persons living with HIV in our cohort was small. Furthermore, we did not have any clinical HIV (e.g., CD4 count) to describe the severity of the disease manifestation. Similar to our findings, a prospective cohort study conducted in Vietnam reported that the hazard rate of post-TB deaths among persons with TB and HIV was 5.1 (95%CI 4.2 – 6.3) times the hazard rate among individuals with TB but without HIV.^25^ However, this hazard estimation combined individuals who died before and after TB treatment. Another study using data from the HIV Epidemiology in South American networks reported a higher hazard rate of death at five years after TB treatment completion among person living with HIV and diagnosed with TB within 30 days of enrollment to the HIV clinic vs. those without TB (aHR 1.57, 95%CI 1.25 – 1.99).^27^ In the same study, the authors reported that among persons with TB and HIV, a lower CD4 count was associated with higher rates of deaths post-TB treatment (aHR 1.57, 95%CI 1.41 – 1.76). Unlike this South America’s study, which only included persons with drug-sensitive TB (DSTB), our study only included individuals with RR and M/XDR TB, and this may partially explain the higher effect of HIV on post-TB mortality reported in our study. Furthermore, our study also had longer follow-up time (up to eight years of follow-up time) and we utilized competing risks model to distinctly modeled the hazard rates of deaths observed during and after TB treatment.

Hyperglycemia has been associated with increased risk of mortality during TB treatment.^28-30^ However, the association between hyperglycemia and the risk of post-TB mortality is not well characterized. We reported a non-significantly higher crude rates as well as the relative effect for post-TB treatment mortality among TB survivors with diabetes compared to those with pre-diabetes or euglycemic. A retrospective cohort study using data from the vital statistics conducted in Israel found that there were less deaths due to diabetes among individuals successfully treated for TB disease compared to the general population.^4^ In contrast, a prospective study conducted in Mexico reported a higher proportion of non-TB related deaths post-TB treatment among individuals with diabetes (4.9%) compared to those without diabetes (2.5%).^31^ The inconsistent findings of the association between pre-existing diabetes and the risk of post-TB deaths may be affected by the use of blood glucose-lowering agents and blood glucose management during and post-TB treatment. For instance, metformin use was associated with reduced mortality risk among individuals with TB in a study conducted in Taiwan.^32^ Prospective clinical studies aiming to assess the impact of diabetes clinical characteristics (including the use of blood glucose-lowering agents during TB treatment) on the risk of mortality post-TB deaths are still warranted.

In our cohort, TB survivors with HCV co-infection had 25% higher rate of post-TB mortality compared to those without HCV co-infection. It is unclear why individuals with HCV who were successfully treated for TB were more likely to die after TB treatment is ended when compared to those with poor TB treatment outcomes. More clinical data (including information whether individuals had acute vs. chronic infections) are needed to understand this finding. To our knowledge, to date, there are limited studies looking at the long-term outcomes of individuals treated for TB with HCV co-infection. It is well known that anti-tuberculosis therapy, especially SLDs, may cause hepatotoxicity and could lead to adverse TB treatment outcomes.^33,34^ Hepatotoxicity could also lead to permanent damage to the liver and may increase the risk of deaths.^35^ Alternatively, post-TB deaths we were observing among individuals with HCV co-infection in this cohort may be due to HCV-related liver disease. However, since information on cause of death as well as time of HCV infection are not known in this cohort, more epidemiologic evidence is needed to link HCV to mortality post-TB treatment.

There are several plausible biological pathways to explain how pre-existing comorbidities could increase the risk of post-TB mortality. First, individuals with pre-existing comorbidities may experience prolonged chronic and systematic inflammation during TB treatment, which could lead to severe disease manifestations/complications (e.g., opportunistic infections among persons living with HIV co-infection, renal or heart disease among individuals with diabetes, and cirrhosis among individuals with HCV co-infection) ^36,37^ which, in turn, will increase the risk of mortality post-TB treatment. Second, individuals successfully treated for TB with pre-existing comorbidities like T2DM may have carry over or residual complications such as proatherogenic and abnormal lipid plasma profile, an established risk factor for cardiovascular diseases.^38^ Third, individuals with pre-existing comorbidities may also have increased risks of TB relapse (i.e., re-infection or recurrent after treatment) due to their impaired immune system.^39-41^ Further clinical studies are needed to establish which of these pathways is more common among TB survivors with pre-existing comorbidities.

Our study subjects to several limitations. First, we performed our analyses among individuals who initiated treatment from 2009 – 2017. Thus, cohort effects associated with changes in national TB treatment guidelines and/or care for individuals with TB and pre-existing comorbidities are likely affected our reported estimates. Second, we did not have data on the cause of death. Thus, we were not able to assess whether mortalities post-TB treatment observed in this study are directly associated with pre-existing comorbidities. Third, we did not have clinical information on key measures for pre-existing comorbidities such as CD4 counts, viral load, blood glucose level during TB treatment, or whether or not individuals were on medications to manage their comorbidities. These important variables may explain how comorbidity factors drove post-TB deaths in this cohort. Thus, future clinical studies including clinical information on the severity of comorbidity factors are still warranted. Fourth, we identified several plausible sources of systematic errors, including bias due to misclassification of hyperglycemia status. However, after running the sensitivity analyses, we did not see any significant difference in our estimates that would change the study’s overall findings. Lastly, our cohort did not include non-TB individuals as a control group. Thus, we were not able to draw direct comparison on mortality rates between individuals with TB vs. non-TB individuals in Georgia. Additionally, our results may not be generalizable to other settings with different TB, HIV, diabetes, and hepatitis C burdens.

In conclusion, we reported an overall high rate of post-TB deaths among individuals who initiated treatment for RR and M/XDR TB, especially among those with pre-existing comorbidities. More importantly, most post-TB deaths were observed within three years after TB treatment was stopped/completed. Aggressive approaches to diagnose comorbidities and integrating TB and non-communicable diseases care to reduce the risk of mortality during TB treatment are needed to help prevent premature deaths. Additionally, continuous care (e.g., linkage to care) for pre-existing comorbidities (especially HIV co-infection) after TB treatment may reduce the increased risks of post-TB deaths.

## Data Availability

All data produced in the present study are available upon reasonable request to the authors.

## ACKNOWLEDGEMENTS

We would like to thank Mamuka Chincharauli for his assistance in pooling data from the Georgia’s TB online surveillance system.

## AUTHOR CONTRIBUTIONS

ADS, MJM, and RRK conceived the study design. ADS and MK obtained and had access to the full data. ADS performed the analyses. ADS, MJM, HMB, RRK, MK provided interpretation of data for the work. ADS wrote the first draft of the manuscript. MJM, HMB, RRK revised the manuscript critically for important intellectual content. All authors reviewed and approved the final version of the manuscript.

## DATA SHARING STATEMENT

Data that support the findings were collected for surveillance purposes and only accessible to investigators upon request to Georgia’s National TB Program and Statistics Office. Individuals included in this cohort did not give written consent for their data to be shared publicly, and since the full data contain sensitive information including individuals’ private health information (e.g., name and date of birth for matching purposes), the full dataset are not made publicly available. However, deidentified individual data, will be made available upon request to corresponding authors (ADS).

## REFERENCES

1 World Health Organization. Global Tuberculosis Report 2022. (World Health Organization, Geneva, 2022).

2 World Health Organization. The END TB strategy. (World Health Organization, Geneva, 2015).

3 Romanowski, K. et al. Long-term all-cause mortality in people treated for tuberculosis: a systematic review and meta-analysis. Lancet Infect Dis, doi:10.1016/S1473-3099(19)30309-3 (2019).

4 Shuldiner, J., Leventhal, A., Chemtob, D. & Mor, Z. Mortality after anti-tuberculosis treatment completion: results of long-term follow-up. Int J Tuberc Lung Dis 20, 43–48, doi:10.5588/ijtld.14.0427 (2016).

5 Harries, A. D. et al. Successfully treated but not fit for purpose: paying attention to chronic lung impairment after TB treatment. Int J Tuberc Lung Dis 20, 1010–1014, doi:10.5588/ijtld.16.0277 (2016).

6 Pearson, F., et al. Tuberculosis and diabetes: bidirectional association in a UK primary care data set. J Epidemiol Community Health 73, 142-147, doi:10.1136/jech-2018-211231 (2019).

7 Salindri, A. D., Wang, J. Y., Lin, H. H. & Magee, M. J. Post-tuberculosis incidence of diabetes, myocardial infarction, and stroke: Retrospective cohort analysis of patients formerly treated for tuberculosis in Taiwan, 2002 - 2013. Int J Infect Dis, doi:10.1016/j.ijid.2019.05.015 (2019).

8 Magee, M. J. et al. Convergence of non-communicable diseases and tuberculosis: a two-way street? Int J Tuberc Lung Dis 22, 1258–1268, doi:10.5588/ijtld.18.0045 (2018).

9 Blondal, K., Rahu, K., Altraja, A., Viiklepp, P. & Rahu, M. Overall and cause-specific mortality among patients with tuberculosis and multidrug-resistant tuberculosis. Int J Tuberc Lung Dis 17, 961–968, doi:10.5588/ijtld.12.0946 (2013).

10 Christensen, A. S., Roed, C., Andersen, P. H., Andersen, A. B. & Obel, N. Long-term mortality in patients with pulmonary and extrapulmonary tuberculosis: a Danish nationwide cohort study. Clin Epidemiol 6, 405–421, doi:10.2147/clep.S65331 (2014).

11 Dangisso, M. H., Woldesemayat, E. M., Datiko, D. G. & Lindtjorn, B. Correction: Long-term outcome of smear-positive tuberculosis patients after initiation and completion of treatment: A ten-year retrospective cohort study. PLoS One 13, e0196432, doi:10.1371/journal.pone.0196432 (2018).

12 Wang, X. H. et al. Survival and associated mortality risk factors among post-treatment pulmonary tubercolosis patients in the northwest of China. Eur Rev Med Pharmacol Sci 19, 2016–2025 (2015).

13 Kolappan, C., Subramani, R., Kumaraswami, V., Santha, T. & Narayanan, P. R. Excess mortality and risk factors for mortality among a cohort of TB patients from rural south India. Int J Tuberc Lung Dis 12, 81–86 (2008).

14 Miller, T. L. et al. Mortality hazard and survival after tuberculosis treatment. Am J Public Health 105, 930–937, doi:10.2105/AJPH.2014.302431 (2015).

15 Cox, H. et al. Tuberculosis recurrence and mortality after successful treatment: impact of drug resistance. PLoS Med 3, e384, doi:10.1371/journal.pmed.0030384 (2006).

16 World Health Organization. Definitions and Reporting Framework for Tuberculosis-2013 Revision. (World Health Organization, Geneva, 2013).

17 Wickham, H. ggplot2: Elegant Graphics for Data Analysis (Springer-Verlag, New York, 2016).

18 R Core Team (2023). R: A language and environment for statistical computing v. 4.2.3 (R Foundation for Statistical Computing, Vienna, Austria, 2023).

19 Austin, P. C., Lee, D. S. & Fine, J. P. Introduction to the Analysis of Survival Data in the Presence of Competing Risks. Circulation 133, 601–609, doi:10.1161/CIRCULATIONAHA.115.017719 (2016).

20 Kleinbaum, D. G. & Klein, M. Survival Analysis: A Self-Learning. Third Edition edn, (Springer, 2011).

21 Greenland, S., Pearl, J. & Robins, J. M. Causal diagrams for epidemiologic research. Epidemiology 10, 37–48 (1999).

22 VanderWeele, T. J. & Ding, P. Sensitivity Analysis in Observational Research: Introducing the E-Value. Ann Intern Med 167, 268–274, doi:10.7326/M16-2607 (2017).

23 Georgia, N. S. O. o. Deaths, <https://www.geostat.ge/en/modules/categories/320/deaths> (2020).

24 Basham, C. A., Karim, M. E., Cook, V. J., Patrick, D. M. & Johnston, J. C. Post-tuberculosis mortality risk among immigrants to British Columbia, Canada, 1985-2015: a time-dependent Cox regression analysis of linked immigration, public health, and vital statistics data. Can J Public Health, doi:10.17269/s41997-020-00345-y (2020).

25 Fox, G. J. et al. Post-treatment Mortality Among Patients With Tuberculosis: A Prospective Cohort Study of 10 964 Patients in Vietnam. Clin Infect Dis 68, 1359–1366, doi:10.1093/cid/ciy665 (2019).

26 Tocque, K., Convrey, R. P., Bellis, M. A., Beeching, N. J. & Davies, P. D. Elevated mortality following diagnosis with a treatable disease: tuberculosis. Int J Tuberc Lung Dis 9, 797–802 (2005).

27 Koenig, S. P. et al. Increased Mortality After Tuberculosis Treatment Completion in Persons Living With Human Immunodeficiency Virus in Latin America. Clin Infect Dis 71, 215–217, doi:10.1093/cid/ciz1032 (2020).

28 Baker, M. A. et al. The impact of diabetes on tuberculosis treatment outcomes: a systematic review. BMC Med 9, 81, doi:10.1186/1741-7015-9-81 (2011).

29 Faurholt-Jepsen, D. et al. Diabetes is a strong predictor of mortality during tuberculosis treatment: a prospective cohort study among tuberculosis patients from Mwanza, Tanzania. Trop. Med. Int. Health 18, 822–829, doi:10.1111/tmi.12120 (2013).

30 Boillat-Blanco, N. et al. Transient Hyperglycemia in Patients With Tuberculosis in Tanzania: Implications for Diabetes Screening Algorithms. J Infect Dis 213, 1163–1172, doi:10.1093/infdis/jiv568 (2016).

31 Jimenez-Corona, M. E. et al. Association of diabetes and tuberculosis: impact on treatment and post-treatment outcomes. Thorax 68, 214–220, doi:10.1136/thoraxjnl-2012-201756 (2013).

32 Degner, N. R., Wang, J. Y., Golub, J. E. & Karakousis, P. C. Metformin Use Reverses the Increased Mortality Associated With Diabetes Mellitus During Tuberculosis Treatment. Clin Infect Dis. 66, 198–205, doi:10.1093/cid/cix819 (2018).

33 Ungo, J. R. et al. Antituberculosis drug-induced hepatotoxicity. The role of hepatitis C virus and the human immunodeficiency virus. Am J Respir Crit Care Med 157, 1871–1876, doi:10.1164/ajrccm.157.6.9711039 (1998).

34 Kempker, R. R. et al. Acquired Drug Resistance in Mycobacterium tuberculosis and Poor Outcomes among Patients with Multidrug-Resistant Tuberculosis. Emerg Infect Dis 21, 992–1001, doi:10.3201/eid2106.141873 (2015).

35 Zhao, H., Wang, Y., Zhang, T., Wang, Q. & Xie, W. Drug-Induced Liver Injury from Anti-Tuberculosis Treatment: A Retrospective Cohort Study. Med Sci Monit 26, e920350, doi:10.12659/MSM.920350 (2020).

36 Kornfeld, H. et al. High Prevalence and Heterogeneity of Diabetes in Patients With TB in South India: A Report from the Effects of Diabetes on Tuberculosis Severity (EDOTS) Study. Chest 149, 1501–1508, doi:10.1016/j.chest.2016.02.675 (2016).

37 Samuels, J. P., Sood, A., Campbell, J. R., Ahmad Khan, F. & Johnston, J. C. Comorbidities and treatment outcomes in multidrug resistant tuberculosis: a systematic review and meta-analysis. Sci Rep 8, 4980, doi:10.1038/s41598-018-23344-z (2018).

38 Vrieling, F. et al. Patients with Concurrent Tuberculosis and Diabetes Have a Pro-Atherogenic Plasma Lipid Profile. EBioMedicine 32, 192–200, doi:10.1016/j.ebiom.2018.05.011 (2018).

39 Dooley, K. E. & Chaisson, R. E. Tuberculosis and diabetes mellitus: convergence of two epidemics. The Lancet Infectious Diseases 9, 737–746, doi:10.1016/S1473-3099(09)70282-8 (2009).

40 Getahun, H., Gunneberg, C., Granich, R. & Nunn, P. HIV infection-associated tuberculosis: the epidemiology and the response. Clin Infect Dis 50 Suppl 3, S201–207, doi:10.1086/651492 (2010).

41 Wu, P.-H., Lin, Y.-T., Hsieh, K.-P., Chuang, H.-Y. & Sheu, C.-C. Hepatitis C Virus Infection Is Associated With an Increased Risk of Active Tuberculosis Disease: A Nationwide Population-Based Study. Medicine 94, e1328–e1328, doi:10.1097/MD.0000000000001328 (2015).

