## Supplementary Table S1 for "HIV co-infection increases the risk of post-tuberculosis mortality among persons who initiated treatment for drug-resistant tuberculosis"

**SUPPLEMENTARY MATERIALS**

|  |  | Page |
| --- | --- | --- |
| **Table S1** | Multiple multivariable models with different covariate specifications  This table was created to show the range of adjusted hazard rate ratios of post-TB mortality from various multivariable models with different set of predictors to account for systematic errors due to covariates misspecifications. | iii |
| **Table S2** | All-cause post-tuberculosis mortality rates among persons who initiated treatment for RR and M/XDR TB in Georgia, 2009 – 2017 (N=998)  This table is the extension of Table 2. In this table, we reported post-TB mortality rates according to other key factors including other comorbidities, smoking status, and baseline body mass index category. | iv |
| **Table S3** | Crude and adjusted hazard rate ratios of mortality during and after tuberculosis treatment among persons who initiated treatment for RR and M/XDR TB in Georgia, 2009 – 2017 (N=1,032)  This table is the extension of Table 3. In this table, we reported crude and adjusted hazard rate ratios of mortality during and after tuberculosis treatment comparing other key factors including other comorbidities, smoking status, and baseline body mass index category. | v |
| **Table S4** | Stratified analyses to assess the association between pre-existing comorbidities and mortality post-tuberculosis treatment according to their final TB treatment outcomes among adult tuberculosis patients previously treated with second-line tuberculosis drugs in Georgia, 2009 – 2017 (N=980)^#^  To better understand about the high-risk group for post-TB mortality, we also ran an additional test where we stratified our primary analysis according to study participants’ final TB treatment outcomes. | vi |
| **Table S5** | Sensitivity and subgroup analyses accounting for systematic errors on the association between various comorbidities and all-cause mortality post-tuberculosis among patients previously treated with second-line tuberculosis drugs in Georgia, 2009 – 2017  We performed a series of sensitivity analyses to quantify the effect of systematic errors on our study estimates. In this table, we reported bias-adjusted estimates from sensitivity analyses to account for errors due to a) model specification, b) misclassification of hyperglycemia/diabetes, and c) effect of unmeasured confounders. | vii - viii |

**Table S1** Multiple multivariable models with different covariate specifications

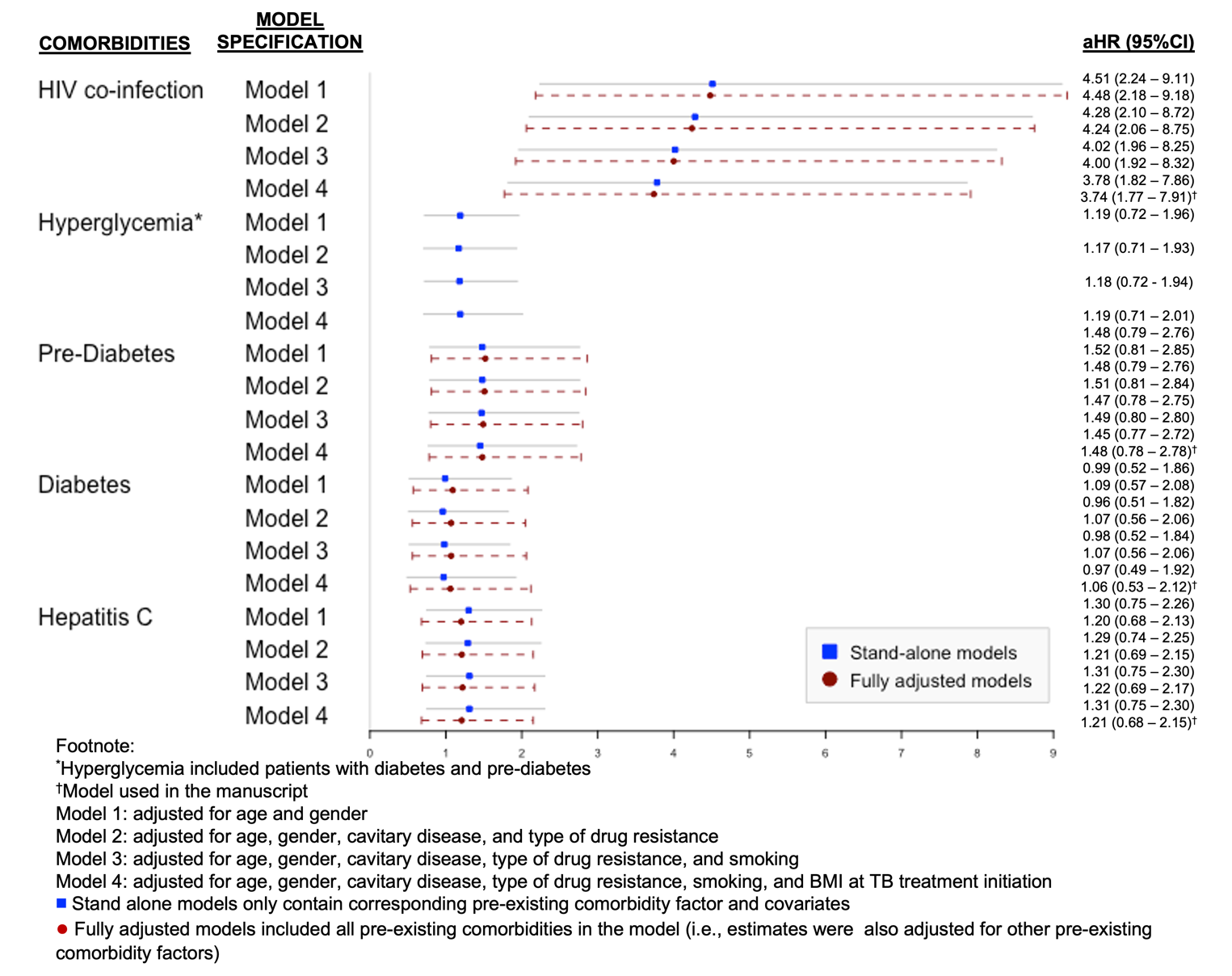

**Table S2** All-cause post-tuberculosis mortality rates among persons who initiated treatment for RR and M/XDR TB in Georgia, 2009 – 2017 (N=998)*

| **Characteristics** | **All-cause mortality post-tuberculosis treatment** | | | | |
| --- | --- | --- | --- | --- | --- |
|  | **Death/Total (%)** | **Person time**  **(Years)** | **Crude Rates (95%CI)**  **per 1,000 person-years** | **Rate Difference (95%CI)**  **per 1,000 person years** | **Median time to death post-TB treatment, months (IQR)** |
| **Any Primary Comorbidity^‡^**  No  Yes | 41/641 (6.4)  46/357 (12.9) | 3128  1729 | 13.11 (9.65 – 17.80)  26.60 (19.93 – 35.52) | Reference  **13.50 (4.83 – 22.17)** | 28 (13 – 41)  19 (6 – 38) |
| **Number of Other Comorbidities^¶^**  0  1 – 2  *Missing* | 60/699 (8.6)  6/52 (11.5)  21/247 (8.5) | 3408  243  1206 | 17.61 (13.67 – 22.67)  24.69 (11.09 – 54.96)  17.41 (11.35 – 26.71) | Reference  7.09 (-13.17, 27.34)  -0.19 (-8.87, 8.49) | 20 (7 – 39)  25 (15 – 28)  28 (7 – 45) |
| ***Other important risk factors*** | | | | | |
| **Smoking status**  Non-smoker  Smoker  *Missing/Unknown* | 10/130 (7.7)  37/437 (8.5)  40/431 (9.3) | 620  2127  2110 | 16.13 (8.68 – 29.98)  17.40 (12.60 – 24.01)  18.96 (13.91 – 25.84) | Reference  1.27 (-10.19, 12.73)  2.83 (-8.77, 14.42) | 32 (15 – 43)  22 (10 – 39)  17 (6 – 39) |
| **Baseline BMI**  Underweight (BMI <18.5)  Normal (BMI 18.5 – 24.9)  Overweight/Obese (BMI ≥25.0)  *Missing* | 16/158 (10.1)  51/633 (8.1)  12/94 (12.8)  8/113 (7.1) | 730  3155  439  533 | 21.92 (13.43 – 35.78)  16.14 (12.29 – 21.27)  27.33 (25.52 – 48.13)  15.01 (7.51 – 30.01) | 5.75 (-5.87, 17.37)  Reference  11.17 (-4.92, 27.26)  -1.16 (-12.46, 10.15) | 20 (5 – 39)  21 (7 – 45)  29 (17 – 36)  7 (4 – 28) |
| **^*^**34 individuals who died during treatment were excluded from the post-TB mortality analyses  ^‡^Any primary comorbidity was defined as the presence of either HIV co-infection, hyperglycemia, or HCV co-infection  **^¶^**Other comorbidities included hepatitis B co-infection, cardiovascular disease, renal disease, ulcer, pancreatitis, and COPD (these were self-reported information and recorded in Georgia’s TB surveillance system)  Abbreviations:  BMI – body mass index; CI – confidence interval; HCV – hepatitis C virus; HIV – human immunodeficiency virus; IQR – interquartile range; TB – tuberculosis | | | | | |

**Table S3** Crude and adjusted hazard rate ratios of mortality during and after tuberculosis treatment among persons who initiated treatment for RR and M/XDR TB in Georgia, 2009 – 2017 (N=1,032)

| **Comorbidities** | **Study Outcomes** | | | |
| --- | --- | --- | --- | --- |
|  | **Tuberculosis mortality during treatment**  **(n=34)** | | **Tuberculosis mortality after treatment**  **(n=87)** | |
|  | **cHR (95%CI)** | **aHR^*^ (95%CI)** | **cHR (95%CI)** | **aHR^*^ (95%CI)** |
| **Any Primary Comorbidity^†^**  No  Yes | Reference  1.42 (0.72 – 2.80) |  | Reference  **2.03 (1.33 – 3.10)** |  |
| **Number of Other Comorbidities^‡^**  0  1 – 2  *Missing* | Reference  1.13 (0.26 – 4.71)  0.95 (0.43 – 2.11) |  | Reference  1.40 (0.60 – 3.23)  0.99 (0.60 – 1.63) |  |
| **Smoking status**  Non-smoker  Smoker  *Missing/Unknown* | Reference  0.91 (0.36 – 2.28)  0.51 (0.19 – 1.41) | Reference  0.85 (0.31 – 2.33)  0.46 (0.16 – 1.31) | Reference  1.07 (0.53 – 2.15)  1.15 (0.58 – 2.31) | Reference  1.00 (0.48 – 2.09)  1.34 (0.66 – 2.74) |
| **Baseline BMI**  Underweight (BMI <18.5)  Normal (BMI 18.5 – 24.9)  Overweight/Obese (BMI ≥25.0)  *Missing* | **3.15 (1.48 – 6.73)**  Reference  1.35 (0.39 – 4.65)  1.49 (0.49 – 4.48) | 2.87 (0.31 – 2.33)  Reference  1.34 (0.34 – 5.20)  1.36 (0.42 – 4.36) | 1.37 (0.78 – 2.41)  Reference  1.72 (0.92 – 3.23)  0.93 (0.44 – 1.97) | 1.31 (0.72 – 2.38)  Reference  1.20 (0.60 – 2.40)  0.88 (0.40 – 1.95) |
| **^*^**Model adjusted for HIV-coinfection, diabetes status, HCV co-infection, age, gender, cavitary disease, type of drug resistance, smoking, and BMI at TB treatment initiation (fully-adjusted model)  ^†^Any comorbidity was defined as the presence of either hyperglycemia, hepatitis C, or HIV co-infection  ^‡^Other comorbidities such as cardiovascular disease, renal disease, ulcer, pancreatitis, COPD, silicosis, sarcoidosis were included  Abbreviations:  aHR – adjusted hazard rate ratios; BMI – body mass index; cHR – crude hazard rate ratios; CI – confidence interval; HCV – hepatitis C virus; HIV – human immunodeficiency virus  **Bold** indicates that the finding is statistically significant at α=0.05  Areas shaded in grey indicate that variables were not included in the final models, and thus, adjusted hazard rate ratios were not obtained. | | | | |

**Table S4** Stratified analyses to assess the association between pre-existing comorbidities and mortality post-tuberculosis treatment according to their final TB treatment outcomes among adult tuberculosis patients previously treated with second-line tuberculosis drugs in Georgia, 2009 – 2017 (N=980)^#^

| **Pre-existing comorbidities** | **aHR^*^ (95%CI)** | |
| --- | --- | --- |
|  | **Among those with favorable^†^ treatment outcomes (n=655)** | **Among those with poor^‡^ treatment outcomes (n=325)** |
| HIV co-infection |  |  |
| Negative | Reference | Reference |
| Positive | 3.30 (0.43 – 25.28) | **3.56 (1.66 – 7.65)** |
| Hyperglycemia |  |  |
| No | Reference | Reference |
| Yes | 1.03 (0.42 – 2.50) | 1.05 (0.57 – 1.94) |
| Diabetes Status |  |  |
| No diabetes | Reference | Reference |
| Pre-diabetes | 1.10 (0.31 – 3.93) | 1.45 (0.69 – 3.07) |
| Diabetes | 0.99 (0.35 – 2.80) | 0.78 (0.35 – 1.75) |
| HCV co-infection |  |  |
| Negative | Reference | Reference |
| Positive | 2.04 (0.81 – 5.13) | 0.99 (0.49 – 2.01) |
| Abbreviations: aHR – adjusted hazard rate ratios; CI – confidence interval; HIV – human immunodeficiency virus  ^#^18 patients with missing final TB treatment outcome were excluded  ^*^Model adjusted for age and gender  **^†^**Including patients who were cured or completed the treatment  ^‡^Including 44 patients who failed the treatment, 247 who were lost to follow up or 34 who died during treatment  **Bold** indicates that the finding is statistically significant at α=0.05  Notes: Among those who were successfully treated for TB disease, 9/26 (34.6%, 95%CI 18.4 – 54.1) died during the first year after completing TB treatment. The median time to post-TB death among those who were successfully treated for TB disease was 24 months (IQR 10 – 46) with median age at time of post-TB death of 57 years (IQR 49 – 66). Among those who had poor TB treatment outcomes, 29/59 (49.2%, 95%CI 36.6 – 61.8). The median time to post-TB death among those who had poor TB treatment outcomes was 20 months (IQR 6 – 39) with median age at time of post-TB death of 45 years (IQR 36 – 63). | | |

**Table S5** Sensitivity and subgroup analyses accounting for systematic errors on the association between various comorbidities and all-cause mortality post-tuberculosis among patients previously treated with second-line tuberculosis drugs in Georgia, 2009 – 2017

| **No** | **Various Bias/Sensitivity Analyses** | **Post-TB treatment Mortality**  **Measure of Association** | **Comorbidities** | **Crude**  **Estimates** | **Adjusted**  **Estimates** |
| --- | --- | --- | --- | --- | --- |
| **1** | **Model Specification** |  |  |  |  |
|  | *Log binomial logistic regression (N=998)* | Risk ratios | HIV co-infection | **3.63 (1.99 – 6.61)** | **3.12 (1.48 – 6.56)**^+^ |
|  |  |  | Hyperglycemia | **1.59 (1.00 – 2.52)** | 1.27 (0.76 – 2.14)^+^ |
|  |  |  | Prediabetes | 1.49 (0.82 – 2.69) | 1.42 (0.76 – 2.67)^+^ |
|  |  |  | Diabetes | 1.70 (0.96 – 3.01) | 1.12 (0.57 – 2.21)^+^ |
|  |  |  | HCV co-infection | 1.54 (0.92 – 2.58) | 1.20 (0.67 – 2.14)^+^ |
| **3** | **Misinformation Bias** |  |  |  |  |
|  | Probabilistic (beta distribution) bias analysis to account for plausible **non-differential** misclassification of hyperglycemia status (1000 iterations) | Median risk ratio  (2.5^th^ – 97.5^th^ percentile) | Systematic Error |  | **1.78 (1.65 – 2.17)** |
|  |  |  | Total Error (random+systematic) |  | **1.84 (1.10 – 2.92)** |
|  | Probabilistic (beta distribution) bias analysis to account for plausible **differential** misclassification of hyperglycemia status (1000 iterations) | Median risk ratio  (2.5^th^ – 97.5^th^ percentile) | Systematic Error |  | **1.73 (1.21 – 2.37)** |
|  |  |  | Total Error (random+systematic) |  | **1.75 (1.01 – 3.00)** |
| **4** | **Unknown Confounders** |  |  |  |  |
|  | Range of plausible true values adjusting for any unknown confounders using the E-value method^*^ | Range of risk ratios | HIV co-infection |  | 0.69 – 2.72 |
|  |  |  | Hyperglycemia |  | 0.30 – 1.19 |
|  |  |  | Pre-diabetes |  | 0.28 – 1.12 |
|  |  |  | Diabetes |  | 0.32 – 1.28 |
|  |  |  | HCV co-infection |  | 0.29 – 1.16 |
|  | E-value (the magnitude of RR_EU_ and RR_UD_) to explain away the observed estimates | Min (RR_EU_, RR_UD_) | HIV co-infection | ≥ 6.72 |  |
|  |  |  | Hyperglycemia | ≥ 2.56 |  |
|  |  |  | Pre-diabetes | ≥ 2.34 |  |
|  |  |  | Diabetes | ≥ 2.79 |  |
|  |  |  | HCV co-infection | ≥ 2.45 |  |
| Footnote:  ^+^Model adjusted for diabetes status, hepatitis C status, HIV-coinfection age, gender, cavitary disease, type of drug resistance, and smoking (fully-adjusted model)  ^*^Simulated with various combination of RR_EU_ 2.00 – 10.00 and RR_UD_ 2.00 – 10.00 resulting in bias factor of 1.3 – 5.3  Abbreviations:  HCV – hepatitis C virus; HIV – human immunodeficiency virus  RR_EU_ – risk ratio estimating the relationship between exposure of interest (i.e., pre-existing comorbidities) and any unknown confounders  RR_UD_ – risk rate ratio estimating the relationship between outcome of interest (i.e., post-TB all-cause mortality) and any unknown confounders | | | | | |
